# Shared decision making and specific informed consent in patients with aortic aneurysms

**DOI:** 10.1101/2021.09.19.21263263

**Authors:** Marcela Juliano Silva Cunha, Marcelo Passos Teivelis, Cynthia de Almeida Mendes, Conrado Dias Pacheco Annicchino Baptistella, Pedro Vasconcelos Henry Sant’Anna, Nelson Wolosker

## Abstract

**Introduction:** Studies show that vascular surgery patients have the desire to participate more actively in their treatment, but that they find it difficult to express themselves. Patients prefer to know all the therapeutic options available, not just those that the surgeon considers appropriate. With this knowledge, patients can begin to choose the most appropriate therapeutic modality for themselves, also becoming responsible for the therapeutic decision. Therefore, the objective of this paper was to analyze the refusal rate of elective aortic aneurysm surgery in asymptomatic patients after the presentation of a detailed IC form followed by a meeting where the patient and his or her family could analyze each item;

**Methods:** Data from 49 patients who had aneurysms and were offered surgical treatment were retrospectively collected and analyzed.

**Results:** After reading the IC and the described meeting, among the 49 patients, 13 (26.5%) refused surgery. We observed that patients who refused surgery had statistically smaller aneurysms than patients who accepted surgery (9% versus 26%).

**Conclusion:** One-quarter of patients who were indicated for elective surgical correction of aortic aneurysms rejected surgery after SDM, which consisted of the presentation of an IC form followed by a clarification meeting for the patient and his or her family to analyze each item. The only factor that significantly influenced a rejection of the procedure was the size of the aneurysm, so patients who rejected surgery had smaller aneurysms than those who accepted surgery.

## Introduction

The shared decision-making (SDM) process is a two-way communication process in which the patient and surgeon collaborate in choosing the appropriate treatment, incorporating patient preferences and values as well as the best scientific evidence. SDM is considered a tool of high ethical and moral standards and it is important not only because it gives the patient the choice of whether to be treated for his or her disease but also because it raises awareness of the different risks that each procedure can bring and the expectations with each choice^1^.

Studies show that vascular surgery patients have the desire to participate more actively in their treatment but that they find it difficult to express themselves. Patients prefer to know all available therapeutic options, not only those that the surgeon considers appropriate.^2^ With this knowledge, patients are able to choose the most appropriate therapeutic modality for themselves (including non-operative approach), becoming responsible for the therapeutic decision as well.

In Brazil and most of the world, the informed consent (IC) for elective surgeries is usually general and standardized for all types of surgery, with some blank spaces to be filled in by the physician with the variability from case to case. It is not customary for a surgeon to apply a specific form to the patient’s disease. The generalized risks, such as bleeding and infection, as well as the risks associated with blood transfusion are usually described for elective procedures. With these documents, patients cannot distinguish between therapeutic alternatives, including nonoperative treatment.

It has been routine in our institution to offer a personalized and detailed document, as part of the SDM process, explaining the patients’ diseases in case of elective vascular surgeries, the proposed surgeries, and all risks and benefits; this document includes a part that must be handwritten by the patient or his or her family in order to demonstrate their understanding of the information. These data are only filled in after a detailed meeting is held among the doctor, the patient and his or her family. These meetings take up to one hour.

The objectives of this study were to analyze the refusal rate for elective aortic aneurysm surgery in asymptomatic patients after the presentation of a detailed IC form followed by a meeting where the patient and his or her family analyze each item; and to identify specific factors of refusal, comparing the epidemiological data, aneurysm characteristics and surgical techniques proposed among patients who accepted the performance of procedures and those who rejected them.

## Methods

The medical records of 49 patients followed by the vascular surgery team of the “Hospital Municipal da Vila Santa Catarina” (between June 2017 and February 2019) with the diagnosis of asymptomatic aorto-iliac aneurysms were retrospectively studied. This study was approved by the Ethics Committee of the Hospital Israelita Albert Einstein (CAAE: CAAE: 13181919.3.0000.0071). Ethical approval was given by the Institutional Review Boards and informed consent was obtained from the included patients. All of the patients were considered fit for surgery after evaluation by the surgical team as well as the clinical team that managed the preoperative evaluation.

In short, IC described the risk of death, paraplegia and spinal cord ischemia, stroke, the risk for colostomy, reoperation due to hernia, limb ischemia / limb amputation, the risk for hemodialysis and the higher risk of re-intervention for endovascular cases.

All patients took home the IC forms, and after sharing them with their family, they returned for a detailed meeting among doctor, family and patient. During this meeting, the document was read in its entirety, and all points were discussed and clarified. There were blank spaces so that the patient or his/her family could handwrite their possible doubts and also their understanding of the proposed surgery and its related risks. A total of 6 physicians (including 2 experienced vascular surgeons and 4 senior residents) participated in these meetings.

The clinical and epidemiological information of patients was collected (age, sex, diagnosis, percentage that the aneurysm exceeded the size by which the particular type of aneurysm would have a formal indication for surgery, proposed/used surgical technique, presence of hypertension, diabetes mellitus, coronary disease, cerebrovascular disease, history of smoking, chronic renal disease and body mass index) as well as the disease for which the surgical treatment was offered and the proposed surgical technique when more than one technique was available.

The indication for surgery in all patients was based on Society of Vascular Surgery and European Society of Vascular Surgery guidelines, and the following diameters were considered as thresholds for different types of aneurysms: abdominal aortic aneurysms of 5.5 cm in men and 5.0 cm in women; thoracic aortic aneurysms diameters of 6 cm; thoracoabdominal aneurysms diameters of 6 cm; and iliac artery aneurysm diameters(common, internal or external) of 3.0 cm^3,4^. In each patient, the percentage above the threshold (PaT) was calculated, which represents the percentage of the aneurysm size that exceeded the minimum diameter in the formal surgical indication. To calculate the PaT, in each patient, the diameter of the artery with aneurysm that exceeded the threshold value was divided by the threshold value of the surgical indication for that specific artery and was multiplied by 100. The decision between open and endovascular techniques took into consideration the materials available for our public service, and always respected the Instructions for Use (IFU).

The patients were divided into two groups: the REJECTED SURGERY group, which was composed of patients who refused the proposed surgical treatment, and the ACCEPTED SURGERY group, which was composed of patients who accepted the proposed surgeries and were then submitted to them.

Initially, the demographic characteristics between the two groups were analyzed, and later, the relationship between the rejection of surgery and PaT was evaluated.

Qualitative characteristics were described using absolute and relative frequencies, and quantitative characteristics were described using summary measurements (mean, standard deviation, median, minimum and maximum). Associations of the qualitative characteristic of the refusal of surgery were verified using Fisher’s exact tests, and quantitative characteristics were compared according to Student’s t-tests. The analyses were performed using SPSS for Windows software version 22.0, and the tests were performed with a significance level of 5%.

## Results

After reading the IC form and after the described meeting, among the 49 patients, 13 (26.5%) refused surgery. These patients were referred to primary care, with orientation to return to our service if they changed their opinion or if there were any new doubts. Unlike what we expected, the type of surgery proposed, open or endovascular, did not seem to directly influence this decision. We did not offer endovascular approach for patients that refused open repair, since one of the motivations for primarily offering open repair for some patients was the aortic anatomy and the non-adequation to the IFU.

The demographic data and data regarding the type of surgery indicated for both groups are presented in table I. We observed that there was no significant difference between the groups, that most of the patients were male, smokers and with hypertension. Only 2% of the cases were thoracic isolated aneurysms, 4% thoracoabdominal, 15% were transrenal aneurysms and 51% were infrarenal abdominal aortic aneurysms. The type of aortic aneurysm did not significantly influence the decision. Regarding the proposed surgical technique, 61.5% of the patients who did not accept surgical treatment received the proposal of an open repair. Among the patients who chose to undergo surgery, 55.5% were operated on using open repair, and there were no statistically significant differences.

**Table 1.** Qualitative and quantitative analyses in each group

|  | Rejected Surgery |  | Accepted Surgery |  | Total | p value |
| --- | --- | --- | --- | --- | --- | --- |
|  | n = 13 | % | n = 36 | % |  |  |
| **Age** |  |  |  |  |  |  |
| Mean±SD | 70,69±6,44 |  | 72,92±6,85 |  |  | 0,298† |

| BMI |  |  |  |  |  |  |
| --- | --- | --- | --- | --- | --- | --- |
| Mean±SD | 26,41±3,76 |  | 25,73±4,73 |  |  | 0,604† |
| Gender |  |  |  |  |  |  |
| Female | 4 | 30,8 | 13 | 36,1 | 17 | >0,999* |
| Male | 9 | 69,2 | 23 | 63,9 | 32 |  |
| Hypertension |  |  |  |  |  |  |
| Yes | 10 | 76,9 | 30 | 83,3 | 40 | 0,683* |
| Diabetes |  |  |  |  |  |  |
| Yes | 2 | 15,4 | 12 | 33,3 | 14 | 0,297* |
| Myocardical Infarction |  |  |  |  |  |  |
| Yes | 3 | 23,1 | 7 | 19,4 | 10 | >0,999* |
| Stroke/Transient Cerebral Ischemia |  |  |  |  |  |  |
| Yes | 2 | 15,4 | 4 | 11,1 | 6 | 0,65* |
| Smoking |  |  |  |  |  |  |
| Yes | 11 | 84,6 | 31 | 86,1 | 42 | >0,999* |
| POCD |  |  |  |  |  |  |
| Yes | 2 | 15,4 | 10 | 27,8 | 12 | 0,474* |
| CKD |  |  |  |  |  |  |
| Yes | 2 | 15,4 | 9 | 25,0 | 11 | 0,703* |
| Surgical technique |  |  |  |  |  |  |
| Open | 8 | 61,5 | 20 | 55,6 | 28 | 0,755* |
| Endovasc. | 5 | 38,5 | 16 | 44,4 | 21 |  |
| Total | 13 | 26,5 | 36 | 73,5 | 49 |  |

\*Fisher's exact test
†Student's t-test

The qualitative and quantitative analyses for each group is described in table I. We observed that patients who refused surgery had statistically smaller aneurysms (9% above the threshold versus 26% above the threshold). (Table II). Therefore, the only factor that significantly influenced patient’s decision to accept or not the proposed surgery was the size of the aneurysm, meaning that the smaller the aneurysm was, the higher the rate of surgical refusal.

**Table 2.** Percentage above the threshold in patients who refused and who accepted surgery

|  | **Refusal** | **Mean** | **SD** | **Median** | **Min.** | **Max.** | **N** | **p value** |
| --- | --- | --- | --- | --- | --- | --- | --- | --- |
| PaT | No | 0,26 | 0,24 | 0,17 | 0 | 0,94 | 36 | **0,003** |
|  | Yes | 0,09 | 0,11 | 0,05 | 0 | 0,32 | 13 |  |

Student's t-test

## Discussion

There are many different ways that vascular surgeons can describe the risk of aneurysm to patients. Aneurysms can be described as having minimal risk, neglecting it, or may be described as ticking time bombs. A Dutch study from 2015 interviewed 10 men with aneurysms with diameters between 35 and 49 mm. Almost all patients were able to explain the basic concept of aneurysms, but several had conceptual doubts. The authors saw that the patients felt comfortable with the surveillance of small aneurysms, but they observed a size of 55mm to represent an abrupt transition when the aneurysm suddenly became dangerous. ^5^ It is noted that the majority of patients had a low level of education, but this low level was not defined by the author. All patients trusted the vascular surgeon to know what was best for them and considered them the most important source of information.^5^

Similarly to Tomee et al,^5^ in our clinical practice, we had the perception that patients with relatively smaller aneurysms were more prone to refuse surgery due to a bigger fear of complications. As we did not find any studies that analyzed the rates of refusal for surgical treatment in patients with aortic aneurysms after the introduction of instruments that would assist with the SDM or after the implementation of a detailed IC form, we felt motivated to analyze the profile of patients who refused aortic surgery in our service.

Currently, some patients with aneurysms are inconsistently informed about their disease and the available treatments. The amount of information provided is less than what is legally owed. Informing the patient about the available treatment options is an ethical and moral issue. The best option for raising this acknowledgment may not be with technological assistance, such as letting the patient watch or read informative materials about the disease using machines, computerized videos or other impersonal ways. It is preferable to use human resources since there is a professional available at the doctor’s office to give personalized and sensitized information to these patients.^6^

From our point of view, the reason why patients with aneurysms refused to undergo surgical repair is related to the understanding of the information exposed and discussed by our team. These asymptomatic patients consider the risk of death small when they are not operated and accept this risk better than possible surgical complications, which for them would probably have a severe impact on their quality of life. For them, the aneurysm was not exactly a ticking time bomb.

SDM prevents excessive treatment and potentially reduces costs (or at least allows resources to be spent only on patients who truly want operative treatment). In addition to the costs of the procedure itself (in our country, the endovascular technique is more expensive), there are costs associated with follow-ups, exams and re-operations. Aneurysm repair increases health care costs, whether public or private.^7^. On the other hand, some of the patients who refuse surgery may develop aneurysm rupture, and when they are able to reach the hospital, emergency treatment will be more expensive than elective treatment.

Patients with less education, more anxiety and a worse prognosis are more prone to prefer less patient-centered care.^8^ From the available national demographic data, it is known that only 6.6% of Brazilian public health users have higher education levels, 31.1% have completed high school, 54.8% have only completed elementary school, and 7.5% are illiterate. ^9^ It is a limitation that our study did not evaluate the education of our patients but there is an important language barrier in meetings in our population. We believe that the information was better absorbed by the patient and his or her family in written form than when it was only verbally expressed, which might have influenced the increased refusal number.

We consider the relatively small number of patients as a limitation of this study, and our findings may be specific to the Brazilian population, whose socioeconomic and educational characteristics may impact SDM. Additionally, because the study was conducted in a public service hospital with limited resources, complex endovascular techniques (such as fenestrated or branched prostheses, or even the use of endoanchors) were not financially viable options; thus, patients with more complex anatomy were offered only the possibility of open repair, which possibly brings with it the perception of greater risks. Since our study was not randomized, we cannot afirm that this refusal rate would not be the same in these patients without this specific IC.

## Conclusion

In conclusion, one-quarter of patients who were indicated for elective repair of aortic aneurysms rejected surgery after SDM, which consisted in the presentation of an IC form followed by a clarification meeting for the patient and his or her family to analyze each item. The only factor that significantly influenced a rejecting of the procedure was the size of the aneurysm, so patients who rejected surgery in general had smaller aneurysms than those who accept surgery.

## Data Availability

The datasets generated during and/or analysed during the current study are available from the corresponding author on reasonable request.

## Acknowledgments

**not applicable**.

